## Supplementary Table descriptions for "Efficient candidate drug target discovery through proteogenomics in a Scottish cohort" (from another version of this preprint: version 1, 2024-01-30)

Supplementary Table 1. Comprehensive Genomic and Proteomic Annotations of 505 Significant SNPs.

**HUGO**: The official symbol representing the gene according to the HUGO Gene Nomenclature Committee.

**Uniprot**: The identifier for the gene's corresponding protein entry in the Universal Protein resource database.

**SNP**: The rsid of the single nucleotide polymorphism being analysed in the study.

**Chromosome**: The chromosome number where the SNP of interest is located.

**Position**: The exact genomic location of the SNP on the chromosome based on the Genome Reference Consortium Human Build 37 (GRCh37).

**Individuals included in the study**: The number of individuals whose data passed the Quality Control steps and were included for GWAS of this protein measurement.

**Effect allele**: The allele associated with the change in studied protein level.

**Other allele**: The alternative allele present at the SNP locus not associated with the effect.

**Effect allele frequency in VIKING**: The frequency of the effect allele in the VIKING cohort.

**Effect allele freq in non-Finnish Eur**: The frequency of the effect allele in the non-Finnish European population (gnomAD).

**Effect Size (beta)**: The estimate of the effect size of the allele in terms of the change in studied protein level.

**Standard error of beta**: The standard error associated with the beta estimate.

**P-value**: The statistical significance of the association between the SNP and the trait.

**Variance Explained**: The proportion of phenotypic variance explained by the SNP.

**Somamer targeted protein chromosome**: The chromosome where the protein targeted by the somamer is encoded.

**Somamer targeted protein TSS**: The transcription start site (TSS) of the gene encoding the somamer targeted protein (GRCh37).

**Bp distance between targeted protein TSS and SNP**: The distance in base pairs between the SNP and the TSS of the somamer targeted protein's gene.

**cisTrans**: Indicates whether the SNP effect is in cis (near the gene it affects, <1Mb) or in trans (affecting a gene located elsewhere, >1Mb).

**Overlapped Gene**: The gene that physically overlaps with the SNP locus.

**Nearest Upstream Gene**: The closest gene upstream to the SNP locus.

**Nearest Downstream Gene**: The closest gene downstream to the SNP locus.

**Most severe consequence**: The most critical biological effect associated with the SNP (Variant Effect Predictor, VEP).

**novel**: Indicates whether the measured protein has been present in previous large-scale proteomics panels.

**somamerID**: The identifier for the somamer used in the analysis.

**FlagQC**: Flags for quality control issues or concerns regarding the somamer, both by SomaLogic and inhouse methods.

Supplementary table 2. Catalogue of Proteins Assayed in Previous Large-Scale Proteomics Studies.

**HUGO or Other name**: Symbol representing the gene according to the HUGO Gene Nomenclature or another unclassified name extracted from supplementary large-scale proteomics studies.

**Uniprot**: The identifier for the gene's corresponding protein entry in the Universal Protein resource database.

Supplementary table 3. Detailed Reference of Assayed Proteins with HUGO, Uniprot, and Sequence IDs.

**HUGO**: Symbol representing the gene according to the HUGO Gene Nomenclature

**Uniprot**: The identifier for the gene's corresponding protein entry in the Universal Protein resource database.

**Full Name**: The complete official name of the protein as recognized in biological and proteomic research and provided by SomaLogic.

**Entrez Gene Symbol**: The gene symbol as designated in the Entrez database.

**Sequence ID**: The unique Somamer identifier.

**Somamer ID**: Another unique Somamer identifier provided by Somalogic.

Supplementary table 4. Determining the Medical Relevance of Various Traits and Diseases with a Large Language Model.

**Outcome**: OpenGWAS dataset (ieu-a, ieu-b, ebi-a, ukb-b) outcome names that were used as input for categorisation.

**ChatGPT Pass1-5**: The ChatGPT assignment of whether the trait is medically actionable in each of the 5 passes.

**Prompt**: The prompt used for as the start of input for each of the passes.

Supplementary table 5. Annotated Compilation of openGWAS Datasets.

**id.outcome:** The unique identifier assigned to the particular outcome or endpoint being studied in the dataset.

**trait:** The specific trait or characteristic that is the focus of the study.

**population:** The demographic or ethnic group that the study participants belong to.

**sample_size:** The total number of individuals who participated in the study.

**ncase:** The number of individuals in the study who have the condition or trait being investigated.

**ncontrol:** The number of individuals in the study serving as a control group, without the condition or trait being investigated.

**subcategory:** The separation of outcomes into Binary or Continuous phenotypes.

**consortium**: The name of the consortium or group that carried out the study.

**author**: The primary author or investigator responsible for the study.

**year**: The year the study was conducted or published.

**pmid**: The PubMed identifier for the study.

**outcome**_**name**: The converted name given to the outcome or endpoint studied for ChatGPT annotation.

**Clinically relevant**: A column indicating whether the outcome or trait has significant implications or applications in a clinical setting as assessed with ChatGPT.

Supplementary table 6. Compilation of Significant Bidirectional Mendelian Randomisation Findings.

**MR Exposure**: The HUGO name of the protein used in the Mendelian randomization study to observe its effect on the outcome.

**MR Outcome**: Name of the trait or condition investigated in the Mendelian randomization study.

**Study ID**: The unique identifier assigned to each individual study in OpenGWAS database.

**MR Method**: The specific method utilised in the Mendelian randomization analysis.

**MR Number of SNPs**: The total count of single nucleotide polymorphisms used in the Mendelian Randomisation analysis.

**MR Effect size (beta)**: The magnitude of the effect that the exposure has on the outcome in the Mendelian Randomisation analysis.

**MR Standard Error of beta**: The standard error associated with the effect size, representing uncertainty in the estimate.

**MR p-value**: The p-value in the Mendelian Randomisation analysis, indicating the statistical significance of the observed effect size.

**Reverse MR Number of SNPs**: The total count of single nucleotide polymorphisms used in the reverse Mendelian Randomisation analysis.

**Reverse MR Effect size (beta)**: The magnitude of the effect observed in the reverse Mendelian Randomisation analysis.

**Reverse MR Standard Error of beta**: The standard error associated with the effect size in the reverse Mendelian Randomisation analysis, representing uncertainty in the estimate.

**Reverse MR p-value**: The p-value in the reverse MR analysis, denoting the statistical significance of the observed effect size.

**Reverse MR Method**: The specific method utilised in the reverse Mendelian randomization analysis.

**Supplementary table 7**. Colocalization Analysis Results.

**HUGO**: The gene symbol identifying the protein measured in this study used in the colocalization analysis.

**Uniprot**: The unique identifier from the UniProt database for the protein measured in this study.

**somamerID**: The unique SomaLogic identifier for the SOMAmer (Slow Off-rate Modified Aptamer) used in the analysis to specifically bind to a particular protein.

**Outcome**: The specific trait or health condition that is being studied in colocalization analysis.

**Open GWAS Dataset ID**: The unique OpenGWAS identifier for this dataset.

**Open GWAS Sample size**: The number of samples (case + control) included in the specific open GWAS dataset employed in the analysis.

**Number of SNP**: Count of single nucleotide polymorphisms involved in the colocalization analysis.

**PP.H0.abf**: The posterior probability that none of the datasets have a signal (i.e., a significant association).

**PP.H1.abf**: The posterior probability that the first dataset has a signal, which is not shared with the other dataset.

**PP.H2.abf** The posterior probability that the second dataset has a signal, which is not shared with the other dataset.

**PP.H3.abf**: The posterior probability that there are signals in both datasets, arising from two independent variants.

**PP.H4.abf**: The posterior probability that the datasets share the same signal, arising from the same causal variant.

**Supplementary table 8.** Gene enrichment analysis results.

Gene Ontology Term: Descriptions of Gene Ontology (GO) terms, each accompanied by its specific GO identifier. A combination of associated biological processes, molecular functions, and cellular components, providing an overview of the functional attributes of the genes in the analysed dataset.

Reference Gene Count: Number of genes associated with each GO term within the total reference 20592 Homo sapiens gene set.

Protein match: Number of uploaded proteins that link to a specific GO term.

Expected Protein Count: Number of proteins from the uploaded set that would be associated with each GO term under a random distribution.

Enrichment Direction: Indicates the direction of enrichment for each GO term in the uploaded dataset. A '+' denotes overrepresentation, meaning that the GO term is more prevalent in the dataset than expected based on the reference gene set. Conversely, a '-' signifies underrepresentation.

Fold Enrichment: Ratio that compares the proportion of a specific GO term in the dataset to its proportion in the reference gene set.

Raw P-value: Raw p-values for each GO term in relation to the uploaded dataset, as calculated from Fisher's Exact Test.

FDR: False Discovery Rate (FDR) values for each GO term in relation to the uploaded dataset, providing an estimate of the expected proportion of false positives.
