## Supplementary figures and images for "Efficient candidate drug target discovery through proteogenomics in a Scottish cohort" (from another version of this preprint: version 1, 2024-01-30)

### Supplementary Figure 1 Scree Plot

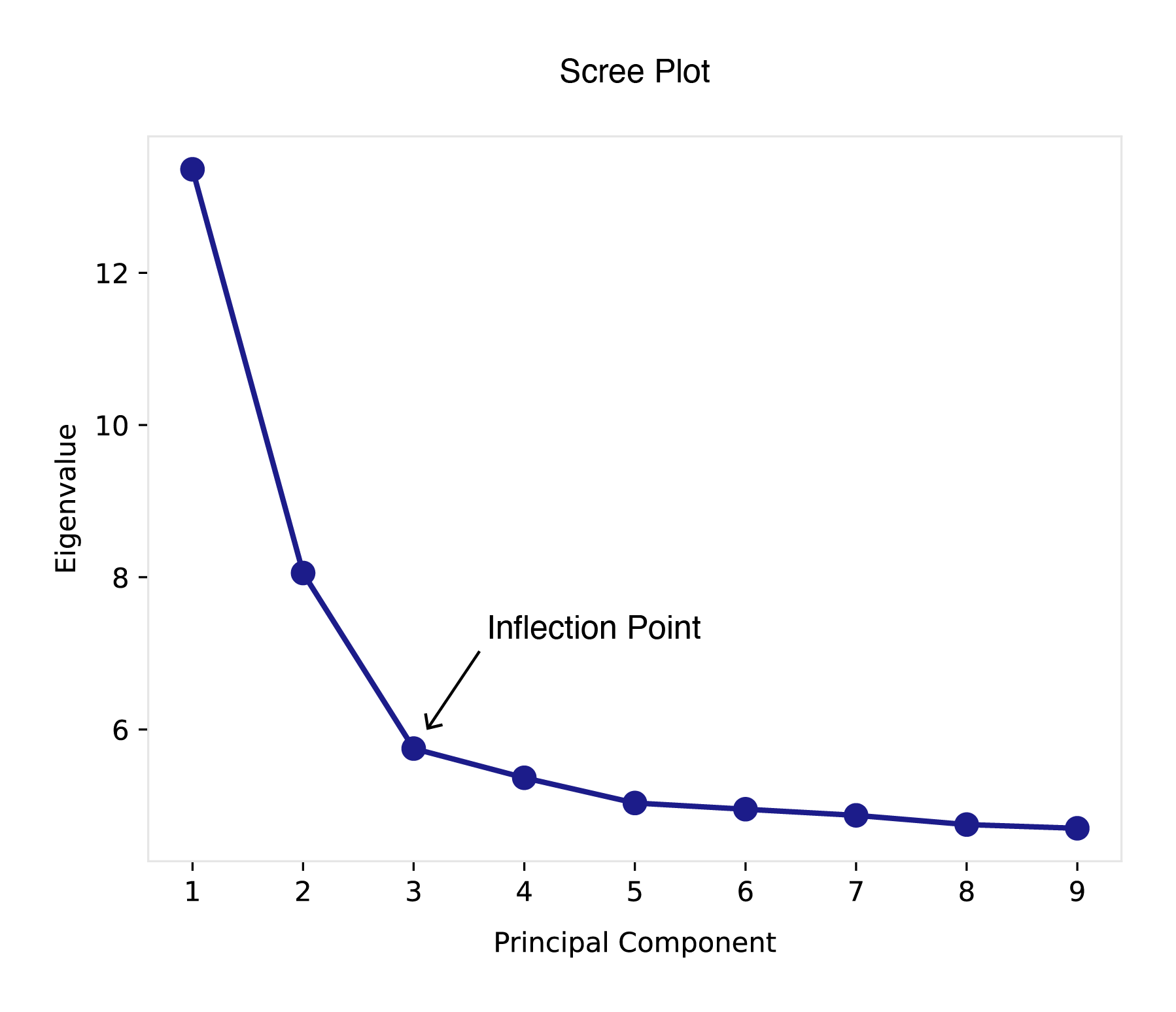

### Supplementary Figure 2 PAV type vs Variance explained

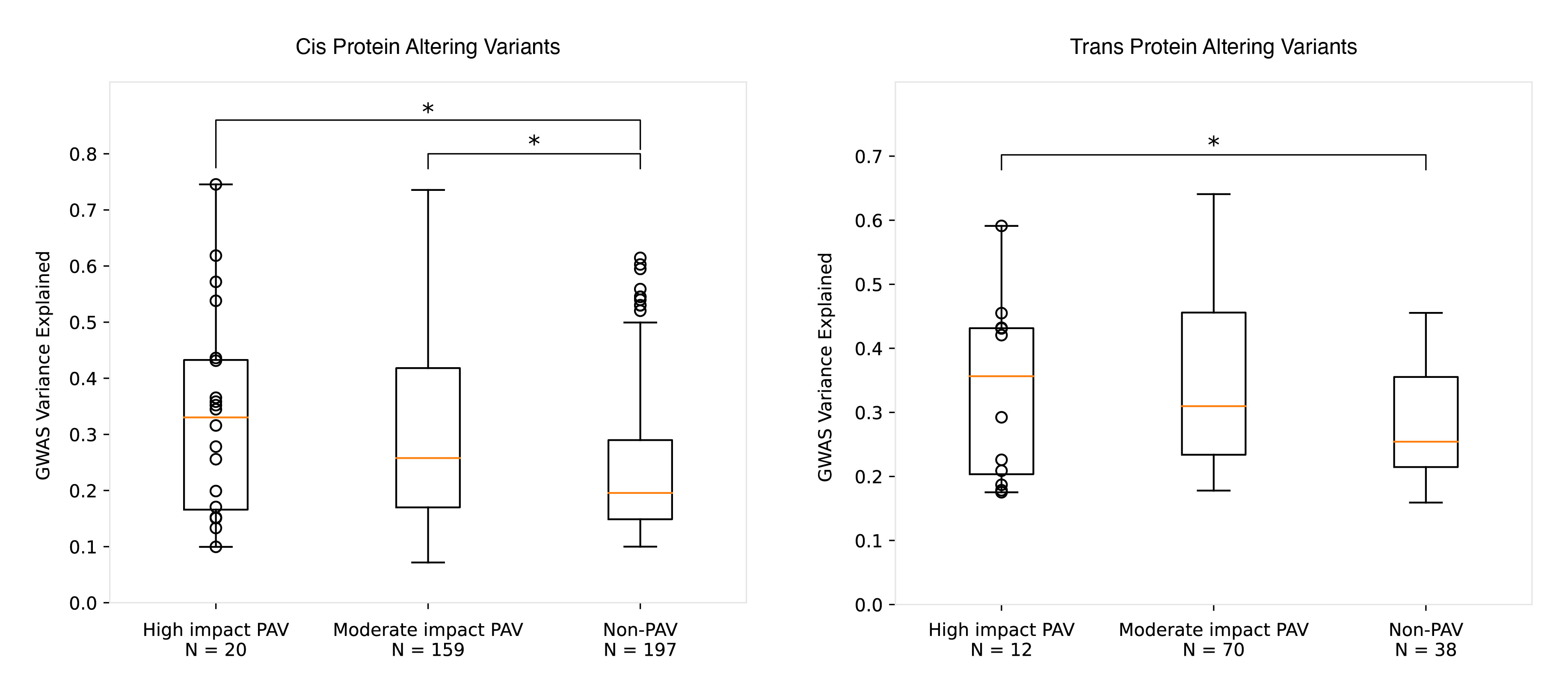

### Supplementary Figure 3 FCGR2B measurement discrepancy

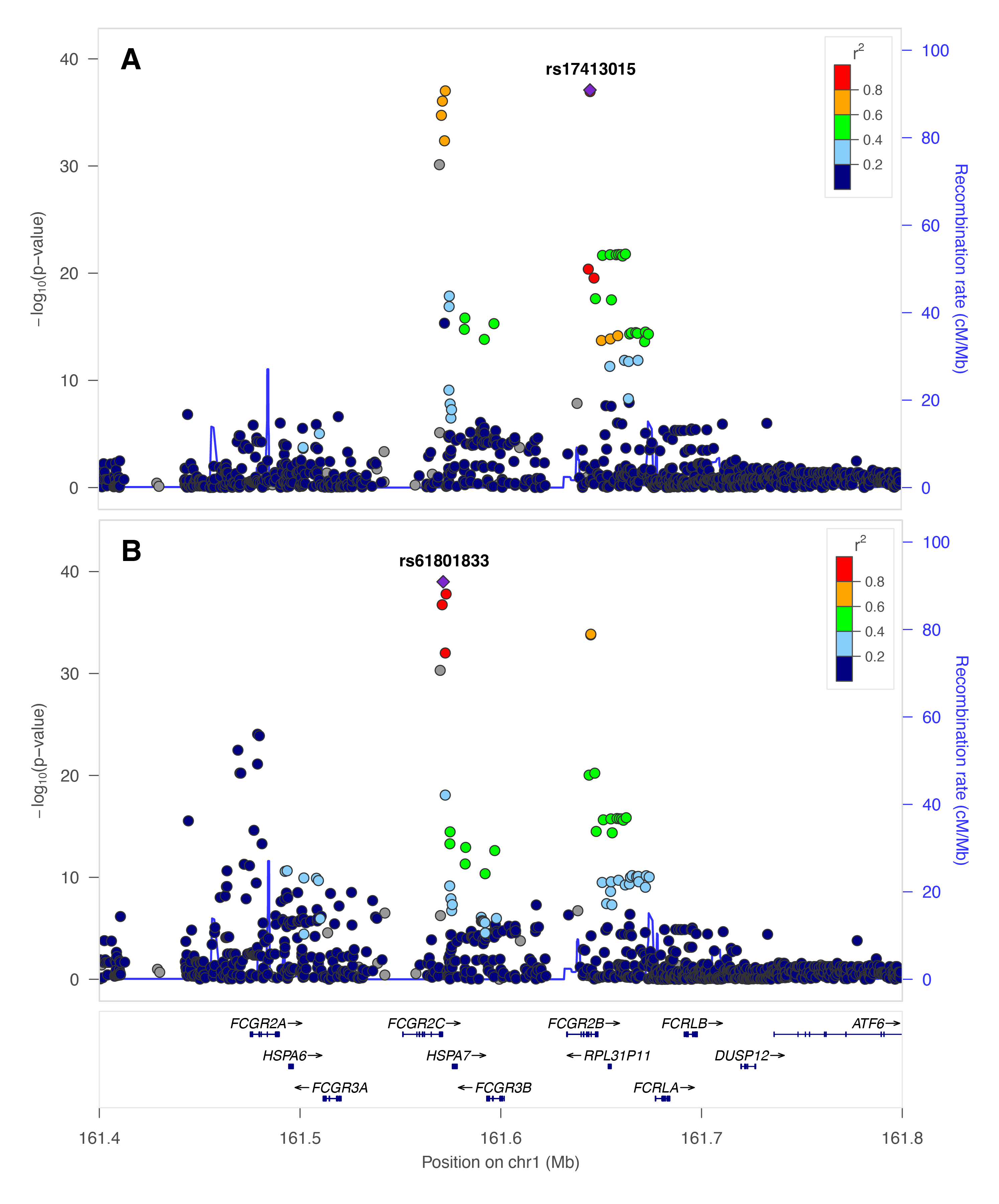
