## Supplementary Figure 4 NIF3L1 LD patterns for "Efficient candidate drug target discovery through proteogenomics in a Scottish cohort" (from another version of this preprint: version 1, 2024-01-30)

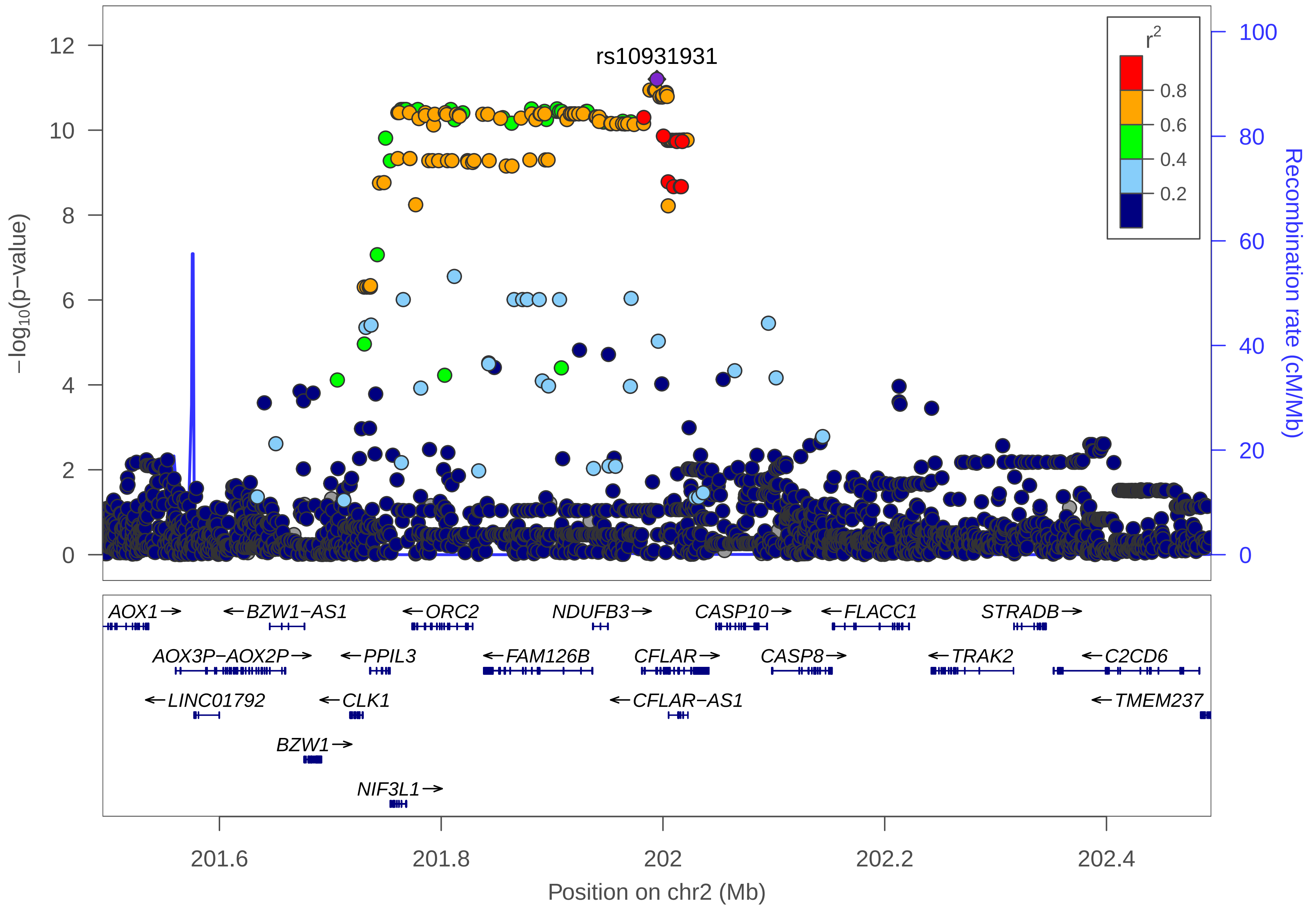
rs10931931
r2
0.8
0.6
0.4
0.2
−log10(p−value)
0
2
4
6
8
10
12
Recombination rate (cM/Mb)
0
20
40
60
80
100
AOX1→
←BZW1−AS1
←ORC2
NDUFB3→
CASP10→
←FLACC1
STRADB→
AOX3P−AOX2P→
←PPIL3
←FAM126B
CFLAR→
CASP8→
←TRAK2
←C2CD6
←LINC01792
←CLK1
←CFLAR−AS1
←TMEM237
BZW1→
NIF3L1→
201.6
201.8
202
202.2
202.4
Position on chr2 (Mb)

| | |
|---|---|
| date: | Wed May 3 19:24:22 2023 |
| build: | hg19 |
| display range: | chr2:201494540−202494540 [201494540−202494540] |
| hilite range: | 0 − 0 [ 0 − 0 ] |
| reference SNP: | chr2:201994540 |
| number of SNPs plotted: | 2351 |
| min p: | 6.3E−12 [chr2:201994540] |
| max p: | 9.95E−1 [chr2:201563844] |

Make more plots at http://csg.sph.umich.edu/locuszoom/
