## Supplementary Figure descriptions for "Efficient candidate drug target discovery through proteogenomics in a Scottish cohort" (from another version of this preprint: version 1, 2024-01-30)

Supplementary Figure 1. Scree Plot Depicting the Relative Contribution of Principal Components to Total Variance. An extended VIKING cohort was used to extract the eigenvalues, comprising 2105, rather than just the 200 individuals with protein level measurements. The plot reveals a steep decline in eigenvalues, with an inflection point observed at PC3, as marked with an arrow. This inflection point delineates the threshold beyond which additional components contribute progressively less to explaining the total variance, underscoring the pivotal role played by the first three components in accounting for the genetic variability in the dataset. It encourages considering PC1, PC2, and PC3 as significant contributors in subsequent analyses, offering a balance between data reduction and retention of information.

Supplementary Figure 2. Protein-Altering Variants (PAV) and Their Contribution to Protein Level Variance. These box plots show that SNPs with a higher impact on protein structure or expression tend to explain a greater variance in the associated protein concentrations compared to non-PAV SNPs. This trend is consistent in both *cis* and *trans* associations. The high-low impact and medium-low impact group differences were statistically significant (p<0.05) for *cis* associations. Meanwhile, for *trans* associations, only the high-low impact group comparison was statistically significant.

Supplementary Figure 3. Aptamer-Specific Differences in FCGR2B Protein Measurement. The showcased LocusZoom plots underline the aptamer-specific differences in capturing the FCGR2B protein measurements, with additional cis-associations in the neighbouring FCGR2A and FCGR2C regions. This differential capture between the two aptamers targeting the same protein shows the lack of specificity in targeting proteins sharing amino acid sequence homology and could influence the resultant variation in protein measurements. A – Aptamer ID 22960-8, B – Aptamer ID 3310-62

Supplementary Figure 4. Linkage Disequilibrium pattern surrounding the NIF3L1 gene. The LocusZoom plot highlights the location of the sentinel variant rs10931931 in relation to NIF3L1 and other nearby genes. The long LD pattern displayed indicate that this region is genetically complex, necessitating further investigation to confidently determine whether rs10931931 is indeed the variant responsible for influencing the risk of age-related macular degeneration through its impact on NIF3L1.
